# Patient-level consequences of local updates to retinal AI for diabetic retinopathy screening: a retrospective two-cohort evaluation

**DOI:** 10.64898/2026.10.23.26366103

**Authors:** Ruiying Li, Teng Ma, Lingtao Tang, Shuman Li, Yiwen Wu, Hong Xia

**Affiliations:** Beijing Tongren Hospital, Capital Medical University, Beijing, China; Department of Ophthalmology, Peking University International Hospital, No. 1 Shengyuan Road, Zhongguancun Life Science Park, Changping District, Beijing 102206, China; School of Biomedical Engineering, Capital Medical University, Beijing 100069, China

**Keywords:** Artificial Intelligence, Diabetic Retinopathy, Mass Screening, Calibration, Machine Learning, Clinical Decision-Making

## Abstract

**Objectives:** To assess whether local artificial intelligence updates for diabetic retinopathy screening that improve image-level probabilities also improve patient-level detection, flagging and net benefit.

**Methods:** This retrospective evaluation used four systems, BRSET source data and two external cohorts, mBRSET and DeepDRiD, with separate audit and evaluation patients. Updates were fitted and selected on audits of 25–200 patients; probability recalibration and operating policies were selected separately. Outcomes included Brier loss, patient detection, flags and net benefit at four threshold probabilities (p=0.05–0.30). Comparators were no update, flag all and flag none. Conditional patient-cluster intervals compared updates with no update; simple-policy comparisons were descriptive.

**Results:** At audit budget 200, mean Brier loss improved in all eight cohort–system comparisons. In mBRSET, flags rose from 154–442 to 809–951 per 1,000 patients and missed positives fell from 51–133 to 2–14 (5.69–9.48 additional flags per additional detection). In mBRSET at p=0.05, three of four updates had lower mean net benefit than flag all; at p=0.20, all four exceeded flag all but remained below no update. In DeepDRiD at p=0.20, the APTOS update increased mean net benefit by 0.0510 over no update and exceeded both simple policies.

**Discussion:** Image-level improvement did not ensure patient-level benefit. Flag increases were consistent with image-level targets applied to patients with about four images.

**Conclusion:** Local updates should be judged on patient-level detection, flags and net benefit against unchanged, flag-all and flag-none policies, not on image-level metrics alone.

**KEY MESSAGES:** *What is already known on this topic:*

- Retinal AI performance can change between settings, and local recalibration or threshold adjustment can improve conventional evaluation metrics.

*What this study adds:*

- Local updates improved image-level Brier loss in all eight cohort–system comparisons, but in mBRSET at p=0.20 all four lowered mean patient-level net benefit, adding 5.7–9.5 positive flags per additional detection. Whether an update outperformed the unchanged, flag-all and flag-none policies depended on the cohort and threshold probability.

*How this study might affect research, practice or policy:*

- Before adopting a local update, services can compare patient detection, positive flags and net benefit with the unchanged, flag-all and flag-none policies on held-out patients.

## INTRODUCTION

Image-based artificial intelligence (AI) systems for diabetic retinopathy screening have shown high diagnostic accuracy, and implementation studies show that their value also depends on the screening pathway in which they operate.[1–3] When a system is used in a different population or acquisition setting, its performance can shift,[4] and recalibration or threshold adjustment may improve it.[5,6] The maintenance and retraining of clinical AI is now a research question in its own right.[7–9]

An update changes decisions as well as prediction error. A hospital deciding whether to adopt an update needs to know how many additional patients would receive positive screening flags and how many positive patients would remain unflagged. Calibration and operating-threshold choice need to be evaluated alongside these patient-level consequences.[10,11] Previous updating studies have assessed calibration and discrimination;[5,6] when several images contribute to one patient’s screening result, an image-level improvement need not carry over to the patient-level decision.

We evaluated whether locally selected updates that improved image-level performance also improved patient-level decisions across four retinal AI systems and two external cohorts, comparing detection, positive flags and net benefit with unchanged and simple screening policies. With limited local audit data, we examined whether selection by patient-level net benefit improved on image-based selection, alongside sensitivity analyses of audit budget, operating targets, source supervision and reference or image coverage.

## METHODS

### Study design and datasets

This was an exploratory retrospective study of publicly released retinal datasets. The four-system comparison on common eligible images, the patient-level selector and the secondary controls were specified after results from earlier stages of this study had been seen. Protocol records, input hashes and numerical checks are listed in the reproducibility materials. We report the two target cohorts separately.

BRSET (PhysioNet version 1.0.2 grades) supplied labels for source fitting, calibration and the source operating policies.[12,13] mBRSET (PhysioNet version 1.0) contains portable-camera photographs from Brazil; DeepDRiD contains conventional colour fundus photographs from China.[14–16] The common four-system analysis included 4,884 graded images from 1,287 mBRSET patients and 1,988 images from 499 DeepDRiD patients. In DeepDRiD, one graded image without an APTOS output was excluded; no patient was lost. Table 1 gives the analysis populations; figure 1 shows the study design.

**Table 1.** Source and target analysis populations.

| Dataset and role | Images | Patients | Audit / evaluation patients | Positive evaluation patients |
| --- | --- | --- | --- | --- |
| BRSET: source fitting/calibration | 16,266 | 8,524 | TRAIN/CAL/TEST<br>5,114/1,705/1,705 | Source-defined |
| mBRSET: four-system common pool | 4,884 | 1,287 | 515 / 772 | 179/772 |
| DeepDRiD: four-system common pool | 1,988 | 499 | 200 / 299 | 149/299 |

Source roles are patient-disjoint within a partition. The three source partitions overlap. Target audit/evaluation counts refer to each partition; audit budgets of 25, 100 and 200 were drawn within the audit pool.

**Figure 1.**
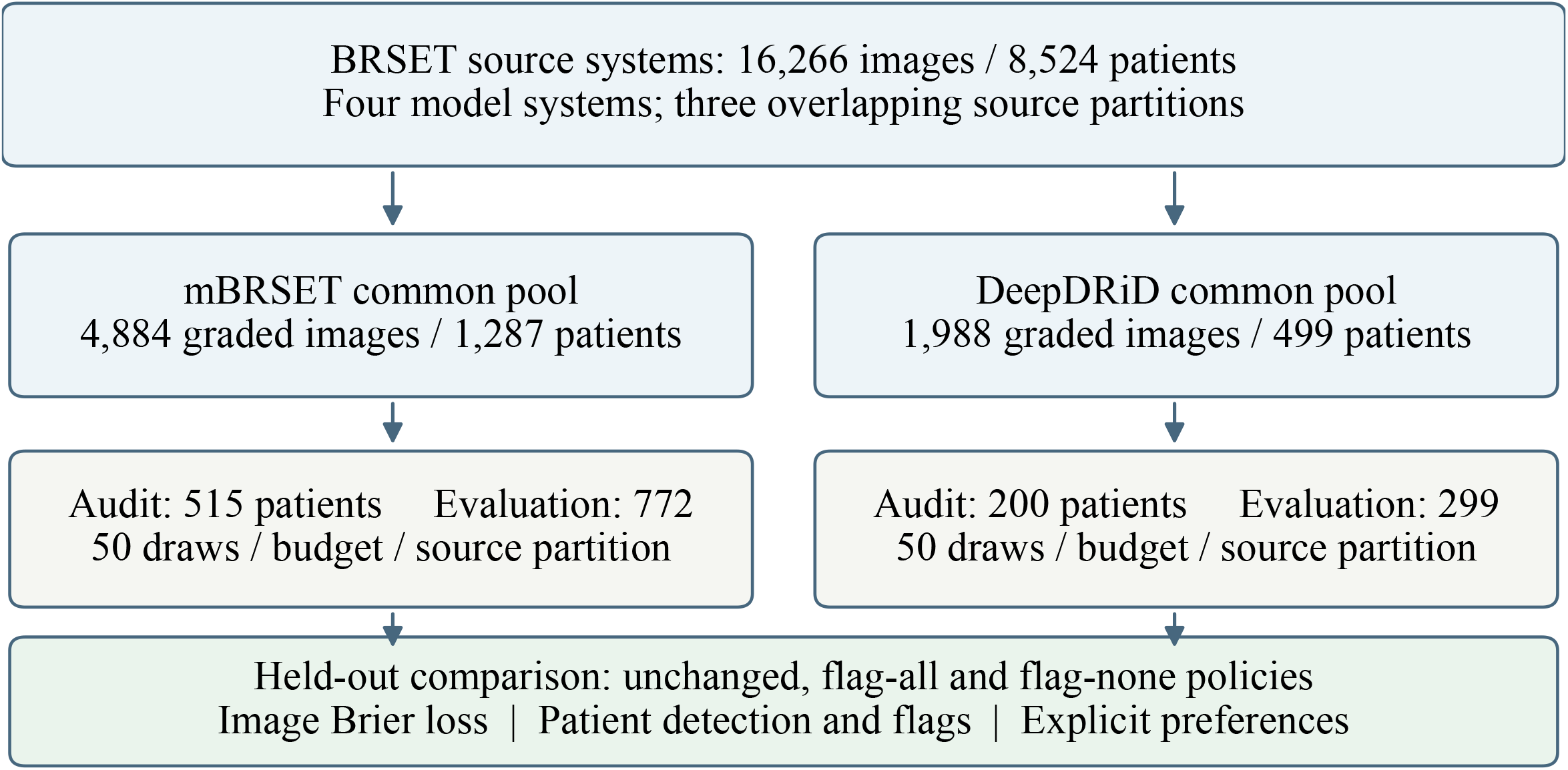
Study design and patient-level assessment. BRSET defined source systems. Shared local audits fitted and selected updates in patient-disjoint target pools; separate patients supplied evaluation outcomes. A patient was flagged when any included image was flagged. Budget 200 used 140 fitting and 60 validation patients. Counts refer to the four-system common-image analysis.

The image endpoint was moderate-or-worse diabetic retinopathy, defined as International Clinical Diabetic Retinopathy grade ≥2 in the released dataset grades. A patient was positive if any included graded image met this endpoint. Missing or ungradable labels were excluded from the labelled image analysis and retained in the eligibility records. A separate sensitivity analysis restricted mBRSET to patients whose recorded images were all graded.

### Systems and local audit procedure

The systems were two general-purpose DINOv3 representations, ViT-S16 and ConvNeXt-T; the retinal foundation representation RETFound; and an author-released, five-fold EfficientNet-B0 ordinal grader trained using APTOS-2019.[17–19] DINOv3 and RETFound used frozen feature extractors with BRSET-trained logistic readouts. The APTOS ensemble’s ordinal score was mapped to a probability using BRSET calibration images. Source calibration and operating thresholds were frozen before each target audit, and the analysis was repeated in three overlapping BRSET source partitions.

Within each source-system partition, patients were separated into local audit and held-out evaluation pools. The pools contained 515 and 772 patients in mBRSET, and 200 and 299 in DeepDRiD. Allocation used patient grades for stratification. Audit budgets were 25, 100 and 200 patients, with 50 shared draws per budget. Each draw was divided into fitting and validation subsets; budget 200 used 140 fitting and 60 validation patients. All systems shared the eligible images and patient draw identities. Evaluation patients contributed to neither update fitting nor action selection.

Candidate actions were no update, intercept recalibration, positive-slope affine recalibration and threshold adjustment.[5,6] The source threshold represented an image-sensitivity target of 0.90. The original operating selector minimised the validation criterion |sensitivity−0.90| + 0.1(1−specificity). For probability estimates, we separately selected no update, intercept or affine recalibration by validation image-level Brier loss. The operating selector determined patient flags, and the independently selected recalibration determined reported probabilities. Audits in which a candidate could not be fitted, or in which no update was selected, remained in the analysis. Detailed recipes and support rules are provided in the supplement.

### Patient-level outcomes and threshold probabilities

A patient received a positive flag when at least one included image crossed the selected cutoff. We calculated patient sensitivity, specificity, missed positive patients and positive flags. The last two quantities were standardised per 1,000 evaluation patients. Outcomes describe screening decisions simulated against released grades.

We assessed the selected decisions with net benefit: NB = TP/N − (FP/N) × p/(1−p), where TP and FP are the numbers of true-positive and false-positive patient flags, N is the number of evaluation patients and p is the threshold probability, so that p/(1−p) weights a false-positive flag against a true-positive detection.[11,20] We used threshold probabilities of 0.05, 0.10, 0.20 and 0.30. These values are illustrative; p=0.20 had been emphasised in an earlier stage of this study. The selected operating policies were held fixed across threshold probabilities and compared with their matched no-update policies. We added flag-all and flag-none reference policies:[21] their net benefits are q−(1−q)p/(1−p) and zero, respectively, where q is the positive patient proportion. We also divided the mean increase in positive flags by the mean reduction in missed positives to express additional flags per additional detection. The ratio includes the newly detected patients among additional flags. A secondary selector chose among the same candidate actions by maximising patient-level validation net benefit at each threshold probability.

### Statistical analysis

We summarised paired action-minus-no-update changes across the 50 audits within each source partition and described their mean across three partitions. Because audits and partitions share patients, these summaries are not independent. Pointwise 95% intervals for Brier and net-benefit differences came from 1,999 bootstrap resamples of evaluation patients within each source partition, conditional on the fixed source system and the saved selected actions. Across-partition means are descriptive. Simple-policy comparisons and additional-flag ratios were added after reviewing the results and calculated descriptively from existing aggregates, without new intervals. Full aggregate estimates and source-specific Brier and net-benefit intervals are supplied in the supplement and accompanying tables; detailed candidate-level support and fallback states are retained in the original analysis records.

Secondary controls included image-sensitivity targets of 0.85 and 0.95 and a source-specific specificity anchor in the two DINOv3 systems. Separate three-representation experiments compared target-only readout replacement with a source-offset readout, and with source thresholds learned using either the validation subset or the full audit. These controls retain their own analysis populations and paired uncertainty. We followed prediction-model and imaging-AI reporting principles in documenting populations, references, model provenance and evaluation units.[22,23]

## RESULTS

### Image-level improvement accompanied substantial patient-level changes

At budget 200, mean Brier loss and the image-level operating criterion improved in all eight cohort–system comparisons. All 24 source-specific conditional intervals for Brier differences were below zero. The patient-level consequences included both improved detection and increased positive flagging (table 2; figure 2).

**Table 2.** Changes in image-level Brier loss and patient-level outcomes at audit budget 200.

| **Cohort / system** | **ΔBrier** | **Flags /1,000:**<br>**unchanged** | **Flags /1,000:**<br>**updated** | **Missed**<br>**/1,000:**<br>**unchanged** | **Missed**<br>**/1,000:**<br>**updated** |
| --- | --- | --- | --- | --- | --- |
| mBRSET / APTOS | -0.0509 | 153.7 | 831.1 | 132.6 | 13.6 |
| mBRSET / DINO-Conv | -0.0280 | 424.0 | 885.5 | 55.3 | 6.6 |
| mBRSET / DINO-ViT | -0.0149 | 441.7 | 809.2 | 51.4 | 10.2 |
| mBRSET / RETFound | -0.0328 | 221.5 | 950.9 | 128.2 | 2.2 |
| DeepDRiD / APTOS | -0.2133 | 427.0 | 800.6 | 132.7 | 17.1 |
| DeepDRiD / DINO-Conv | -0.1630 | 545.2 | 858.3 | 68.0 | 2.3 |
| DeepDRiD / DINO-ViT | -0.0966 | 637.7 | 773.0 | 33.4 | 9.7 |
| DeepDRiD / RETFound | -0.1117 | 622.1 | 871.1 | 54.6 | 8.3 |

Negative $\Delta$ Brier favours updating. Brier loss evaluates the separately selected probability recalibration; detection and flagging evaluate the selected operating policy. DINO-Conv denotes DINOv3 ConvNeXt-T; DINO-ViT denotes DINOv3 ViT-S16. “Missed” denotes positive patients without a positive flag. Means cover three source partitions × 50 dependent audits. Brier is image-level; rates are patient-level. Across-partition means are descriptive; source-specific conditional Brier and net-benefit intervals are in supplementary table S2.

**Figure 2.**
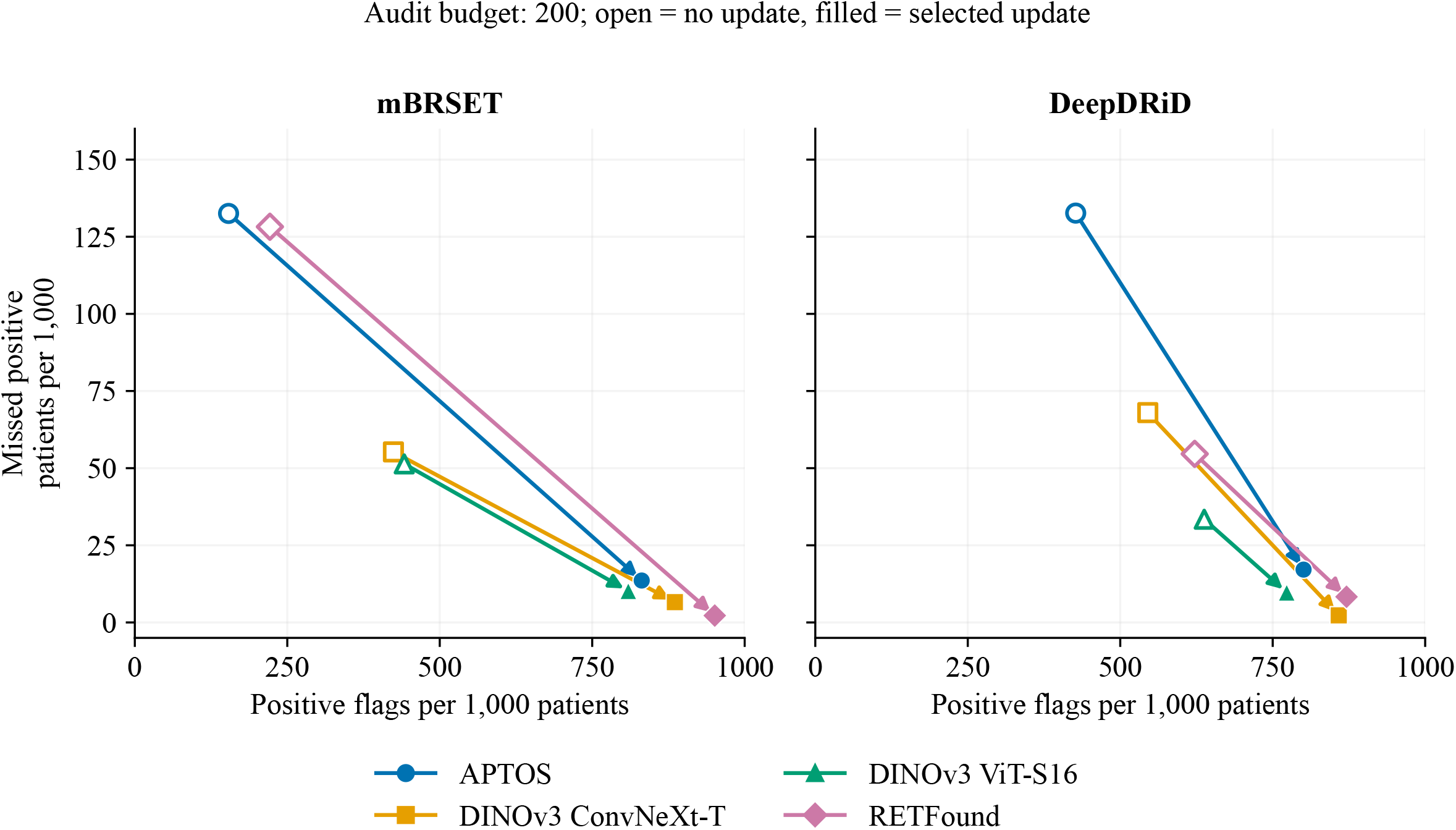
Positive flags and missed positive patients before and after local updating. Arrows connect no update to the selected operating policy at audit budget 200. Each point is a mean across three overlapping source-system partitions and 50 dependent audits per partition; denominators are 772 evaluation patients per partition for mBRSET and 299 for DeepDRiD. Axes show rates per 1,000 evaluation patients. Source-specific flag and miss estimates, and conditional intervals for Brier and net-benefit differences, are supplied in the supplement.

In mBRSET, the APTOS system’s mean flags increased from 153.7 to 831.1 per 1,000 patients, while missed positive patients decreased from 132.6 to 13.6. The corresponding values were 424.0 to 885.5 flags and 55.3 to 6.6 omissions for DINOv3 ConvNeXt-T; 441.7 to 809.2 flags and 51.4 to 10.2 omissions for DINOv3 ViT-S16; and 221.5 to 950.9 flags and 128.2 to 2.2 omissions for RETFound. Updated patient sensitivity ranged from 0.941 to 0.990, with specificity from 0.061 to 0.235. Additional flags per additional positive detection were 5.69, 9.48, 8.92 and 5.79 for APTOS, ConvNeXt-T, ViT-S16 and RETFound, respectively (supplementary table S3).

DeepDRiD had a higher positive proportion: 149/299 evaluation patients, compared with 179/772 in mBRSET. Its mean flag increases ranged from 135.3 to 373.6 per 1,000 patients, alongside omission reductions of 23.7–115.5 per 1,000. Updated sensitivity ranged from 0.966 to 0.995 and specificity from 0.240 to 0.433. The increases in detection and positive flags therefore differed between cohorts and systems.

### The threshold probability changed the assessment of the same policies

Figure 3 compares absolute patient net benefit with the unchanged, flag-all and flag-none policies across threshold probabilities. In mBRSET, all four systems had positive mean net-benefit differences against no update at p=0.05, ranging from 0.0240 to 0.0943. At p=0.20, all four means were negative, ranging from −0.0545 to −0.0206. At this threshold, eight of twelve source-specific conditional intervals were below zero and four included zero. Both DINOv3 systems had negative intervals in all three source partitions; the direction persisted in the fully graded sensitivity subsets. At p=0.05, flag-all net benefit was 0.1914. Selected-update means were 0.1860, 0.1905, 0.1908 and 0.1917 for APTOS, ConvNeXt-T, ViT-S16 and RETFound: three were below flag all, with RETFound only 0.0002 above it. At p=0.20, all four updates exceeded flag all (0.0398) and flag none (zero), but remained below their matched unchanged policies. At p=0.30, all four selected-update means were below flag none, whereas unchanged-policy means remained positive. The comparison therefore depended on both the threshold probability and the reference policy. For illustration, the mBRSET ConvNeXt-T source-partition-0 Brier difference was −0.0349 (95% CI −0.0459 to −0.0251), while its net-benefit difference at p=0.20 was −0.0605 (−0.0809 to −0.0394).

**Figure 3.**
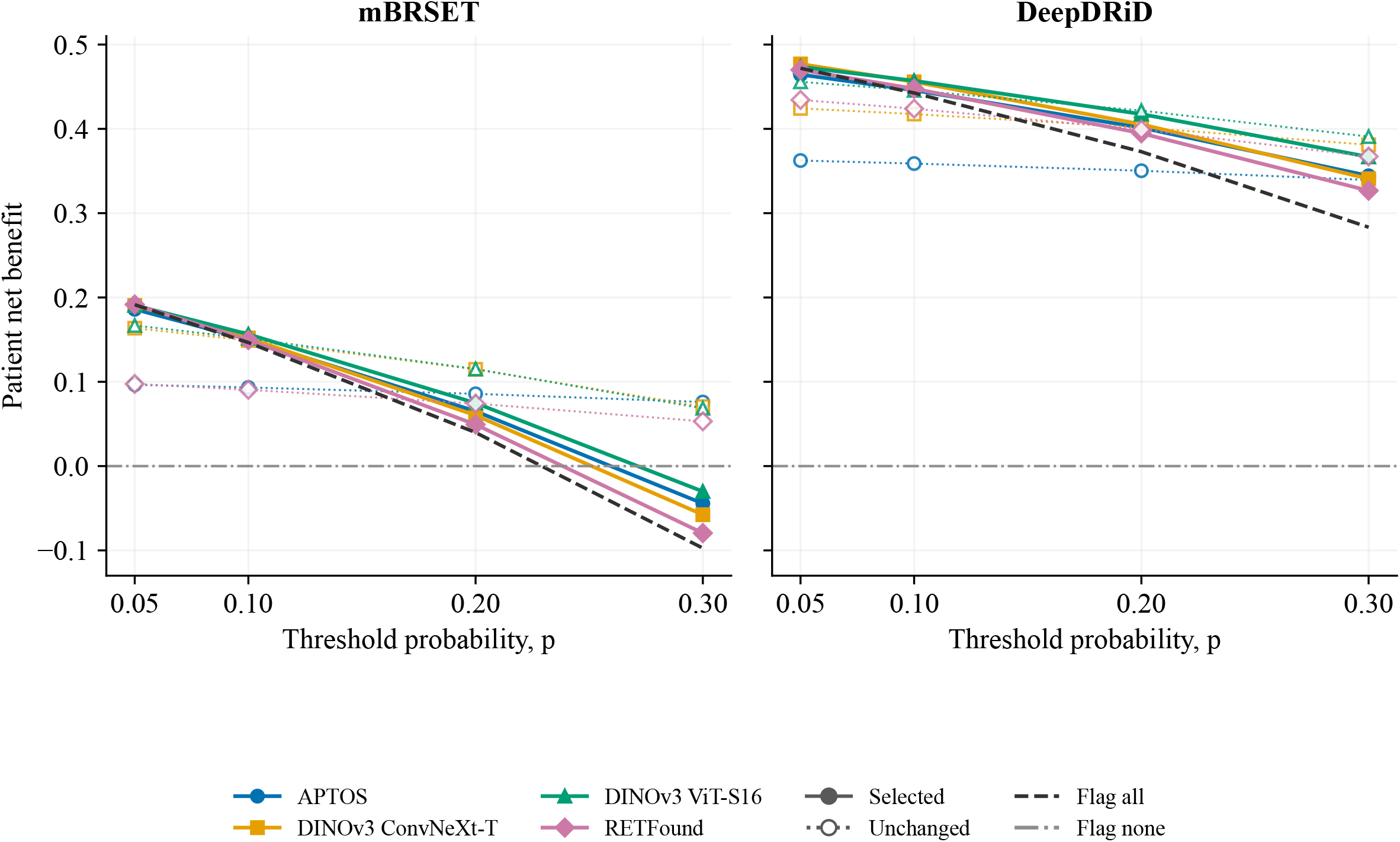
Net benefit of selected and unchanged policies across threshold probabilities. Absolute net benefit is shown at four threshold probabilities. Filled markers and solid lines show selected updates; open markers and dotted lines show matched unchanged policies. Marker shape identifies the system. The dashed line shows flag all and the dash-dot line flag none (zero). Higher values indicate greater net benefit at the stated threshold probability. Points summarise the same three source partitions and 50 audits as figure 2; line segments connect the displayed threshold probabilities while each policy’s decisions remain fixed. Source-specific pointwise conditional intervals for update-minus-no-update differences are given in supplementary table S2. Flag-all/flag-none comparisons are descriptive; supplementary table S3 and the accompanying CSV provide precise means.

The comparison in DeepDRiD was different. At p=0.20, the APTOS update had mean incremental net benefit 0.0510, with all three source-specific intervals above zero. Mean changes for ConvNeXt-T, ViT-S16 and RETFound were 0.0039, −0.0041 and −0.0044; their intervals included zero. The APTOS update also exceeded flag all by a mean 0.0284 and therefore had higher mean net benefit than all three reference policies at this threshold. At p=0.05, however, the APTOS and RETFound updates were 0.0075 and 0.0020 below flag all.

### Selection by patient-level net benefit

At audit budget 200 and p=0.20 in mBRSET, selecting actions by patient-level validation net benefit increased mean net benefit relative to the original image-based selector by 0.0140 for APTOS, 0.0520 for ConvNeXt-T, 0.0388 for ViT-S16 and 0.0211 for RETFound. Mean differences against no update remained negative (−0.0066, −0.0025, −0.0016 and −0.0038), and the selector retained no update in 65–96% of audits. In DeepDRiD at p=0.20, the corresponding changes relative to image-based selection ranged from −0.0057 to 0.0025. Results for other audit sizes and threshold probabilities are provided in the supplement.

### Source supervision and alternative operating targets

In the separate readout experiments, retaining the source-supervised score improved the patient-level area under the receiver operating characteristic curve (AUROC) relative to the frozen source in five of six cohort–representation comparisons at budget 200, with paired intervals above zero. At an audit flagging fraction request of 0.50, paired intervals supported fewer omissions than source thresholds learned from all 200 audit patients in five of six comparisons and more positive flags in four. Against target-only replacement, two AUROC intervals were above zero and four included zero.

Alternative operating targets also changed relative action performance. Among twelve n=200 DINOv3 cohort–system–target comparisons, fixed threshold adjustment had lower mean image operating loss than the audit selector in ten, tied in one and higher loss in one. Reference definitions also changed the assessment of fixed predictions. With images and predictions held fixed, replacing grades from the earlier BRSET release 1.0.0 with those from release 1.0.1 changed sensitivity for the grade ≥3 endpoint by +0.036 and +0.038 in the two DINOv3 systems; all primary analyses used release 1.0.2. In a separate 650-patient mBRSET analysis, two-image reference subsets omitted 11–15 of 151 positive evaluation patients. Complete strategy-specific results appear in the supplement.

## DISCUSSION

Local updates improved image-level Brier loss in all eight cohort–system comparisons, yet patient-level net benefit improved only in some settings. With 200-patient audits, updating increased both patient detection and positive screening flags: in mBRSET, each additional detection was accompanied by 5.69–9.48 additional flags, and in DeepDRiD by 3.23–5.70. Shared images and patient samples across four systems allowed these consequences to be compared directly.

The size of the flagging increase is consistent with how operating policies were chosen. The selector moved each cutoff towards an image-level sensitivity of 0.90, whereas patients contributed about four images each and were flagged when any image crossed the cutoff. Updated patient-level sensitivity reached 0.94–0.99, above the image-level target, and in mBRSET patient specificity fell from 0.66–0.93 to 0.06–0.24. Operating targets for screening updates should therefore be set and checked on the patient-level decision that the service acts on.

The net benefit of an update depended on the receiving cohort, the relative weight assigned to false-positive flags and the comparator. At p=0.05, three mBRSET updates improved on no update but remained below flag all in mean net benefit. At p=0.20, all four exceeded flag all and flag none but remained below the unchanged policy. In DeepDRiD at p=0.20, the APTOS update was the only one whose gain over no update had all three source-specific intervals above zero, and it also exceeded both simple policies. These comparisons show when increased detection offsets the additional positive flags at a stated threshold probability.

Selecting actions by patient-level net benefit, one step in this direction, narrowed the mBRSET deficit at p=0.20 mainly by keeping the unchanged policy. In DeepDRiD, patient-level selection produced a small mean improvement for ViT-S16 and decreases for the other three systems. Because this selector was specified after the image-based results had been seen, it needs testing on new patients. In the separate readout experiments, retaining the source-supervised score improved discrimination and, in several comparisons, reduced missed positives beyond source threshold adjustment. Local audit data can therefore help choose among candidate updates, as closed testing procedures do for regression models,[24] whereas evaluation on separate patients shows whether the chosen update outperforms the existing policy.

These findings extend work on clinical AI maintenance and retraining[7–9] by reporting the patient-level consequences of local updates alongside calibration.[10] Comparing an update with the existing and simple policies gives a service the comparison it needs when deciding whether to replace its current model.

The study was exploratory, using released image grades and patient classifications aggregated across available images. Several comparisons, including the patient-level selector and the simple-policy references, were specified after results from earlier stages of this study had been seen. Some mBRSET records lacked grades; restriction to fully graded records supported the main DINOv3 net-benefit findings. Confidence intervals condition on the observed cohorts, fitted systems and saved selected actions, and the three source partitions share patients. Overlap between target images and the pretraining data of the foundation and third-party models could not be checked. Systems differed in pretraining and adaptation, and the threshold probabilities were illustrative. Actual referrals, resource use and downstream patient outcomes were not measured. Prospective evaluation with adjudicated patient references and service outcomes would establish the clinical value of the observed changes.

## CONCLUSIONS

Local retinal AI updates that improved image-level probabilities increased patient detection but also sharply increased positive screening flags, and their net benefit relative to the existing policy depended on cohort and threshold probability. Updates should be judged on patient-level detection, flags and net benefit against unchanged and simple policies, at an explicit threshold probability.

## Data Availability

Aggregate results, figure source data and publication-reproduction scripts are provided in the supplementary code archive. BRSET and mBRSET are available through PhysioNet credentialed access, and DeepDRiD through its providers, under their respective terms (references 12 to 16). Original images, individual level records, model weights and extracted features are not redistributed. The supplied scripts regenerate the publication tables and figures from these aggregates. The primary analysis and selector scripts are included as source code; rerunning them requires the authorised images, model weights and saved states described in the methods attachment.

https://doi.org/10.13026/mysn-8b26

https://doi.org/10.13026/qxpd-1y65

## DECLARATIONS

### Ethics approval and consent

The authors considered additional ethics approval unnecessary for this secondary analysis of previously released de-identified datasets, which involved no new recruitment, contact or intervention. The source studies’ approval and consent arrangements are documented in the cited provider reports.

### Patient and public involvement

Patients and the public were not involved in the design, conduct, reporting or dissemination planning of this secondary analysis.

### Contributorship and guarantor

Ruiying Li contributed to conceptualisation, data curation, formal analysis, software, visualisation and writing the original draft. Teng Ma contributed to clinical interpretation of the results and writing—review and editing. Lingtao Tang contributed to methodology, software validation, engineering review and writing—review and editing. Shuman Li contributed to data curation, including verification of the datasets and organisation of the model data. Yiwen Wu contributed to data curation, including verification of the datasets, verification of the references and checking of the manuscript formatting. Hong Xia contributed to methodology, statistical interpretation, supervision and writing— review and editing. Ruiying Li is the guarantor. All authors agreed to the final manuscript and submission.

### Registration

This study was not registered.

### Data availability statement

Aggregate results, figure source data and publication-reproduction scripts are provided in the supplementary code archive. BRSET and mBRSET are available through PhysioNet credentialed access, and DeepDRiD through its providers, under their respective terms (references 12–16). Original images, individual-level records, model weights and extracted features are not redistributed. The supplied scripts regenerate the publication tables and figures from these aggregates. The primary-analysis and selector scripts are included as source code; rerunning them requires the authorised images, model weights and saved states described in the methods attachment.

### AI assistance

DeepSeek V4 Pro assisted code development and manuscript language polishing. The authors are responsible for verifying the methods, results, references, interpretation and final submitted text.

## Acknowledgements

We thank the investigators and participants whose data were made available through BRSET, mBRSET and DeepDRiD.

