## supplementary_information for "Patient-level consequences of local updates to retinal AI for diabetic retinopathy screening: a retrospective two-cohort evaluation"

### S1. STUDY SCOPE AND ANALYSIS POPULATIONS

This supplement accompanies *Patient-level consequences of local updates to retinal AI for diabetic retinopathy screening: a retrospective two-cohort evaluation*. The primary publication comparison uses the common four-system image intersection in mBRSET and regular-camera DeepDRiD. The analyses below retain their original data populations and purposes. The disease endpoint is recorded moderate-or-worse diabetic retinopathy; macular oedema is outside this endpoint.

BRSET comprised 16,266 images from 8,524 patients, graded according to the PhysioNet version 1.0.2 label file. It supplied source fitting, calibration and threshold selection. Three source partitions were formed by patient, with nominal 60% training, 20% calibration and 20% source testing; the role counts were 5,114, 1,705 and 1,705 patients. Stratification used the patient maximum recorded grade, capped at 3, and dominant camera. Source testing did not determine the preferred target action.

mBRSET comprised 5,164 released images from 1,291 patients; its common labelled action pool included 4,884 images from 1,287 patients. The audit pool contained 515 patients and the evaluation pool 772. Each source partition's evaluation pool contained 179 positive patients. DeepDRiD had 1,989 eligible graded images before four-system matching; one image lacked an eligible APTOS output, leaving 1,988 images and all 499 patients. Its audit pool contained 200 patients and evaluation pool 299, including 149 positive patients. The source-specific evaluation image counts were 2,928/2,917/2,927 for mBRSET and 1,189/1,195/1,194 for DeepDRiD.

A patient's recorded endpoint was positive if any included image had grade ≥2. A positive predicted flag was the OR of image decisions. Patients with incomplete image grading therefore have an endpoint defined by the available graded records. The complete-grade sensitivity populations in mBRSET contained 651, 651 and 654 evaluation patients in the three source partitions. These differ from the separate 650-patient view-coverage experiment described in S7.

Original acquisition settings, reported collection periods and reference-reader procedures are summarised from the cited provider papers in methods/dataset_reference_context.pdf.

### S2. MODEL PROVENANCE AND SOURCE POLICIES

DINOv3 ViT-S16 and ConvNeXt-T used the frozen feature recipes specified in the model-methods attachment. RETFound used the RF-MAE-NOTEBOOK-GAP feature recipe with a logistic readout. Standardisation and logistic readout fitting used BRSET training images only. The retained readout regularisation was C=0.01, with a maximum of 3,000 iterations. Source probability calibration and operating-threshold selection used the calibration role. Source calibration retained its original fitting routine; local maintenance candidates used the positive-slope bounds specified in S3.

The third-party APTOS system comprised five author-released EfficientNet-B0 ordinal regression folds. We used the locked weights, code and inference recipe specified in the original APTOS admission protocol (methods/THIRD_PARTY_APTOS_MODEL_20260930.md in supplementary_code.zip). Fold scores were averaged before source mapping. BRSET adaptation comprised a positive-slope logistic mapping of the frozen ordinal score and threshold selection, both using calibration patients. Source test patients supplied descriptive evaluation.

The companion methods/model_provenance_and_locked_methods.pdf records all four systems' checkpoint identities, recorded SHA256 hashes, actual preprocessing, feature definitions, inference recipes and source mappings. It also distinguishes the historical admission protocol from the later completed execution records.

The DINOv3 model documentation identifies general web pretraining. The RETFound publication describes retinal self-supervised pretraining including EyePACS. The APTOS model author identifies APTOS-2019 as the training source. Image-level overlap between the target cohorts and these pretraining sets could not be checked because the full pretraining inventories were unavailable. Both target cohorts had been analysed in earlier, unpublished stages of this study, so the maintenance analyses are retrospective extensions rather than first analyses of these cohorts.

### S3. AUDIT FITTING, SELECTION AND NUMERICAL DEFINITIONS

Audit budgets 25, 100 and 200 were divided into 17/8, 70/30 and 140/60 fitting/validation patients. There were 50 saved draws per source partition and budget. Image keys, patient identities, audit roles and eligibility were shared across the four systems. All three source partitions were retained.

No update retained the source score and threshold. Intercept recalibration shifted the source logit; affine recalibration estimated an intercept and a positive slope. Their fitting support required at least two positive and two negative image events. The local affine solver used intercept bounds −100 to 100 and slope bounds 1e−8 to 50, with L-BFGS-B, maximum 500 iterations, ftol 1e−12 and gtol 1e−8. Failed or inadmissible fits remained unavailable. Threshold relocation required at least ten distinct recorded-positive fitting patients. The cutoff was the descending positive-image score at index ceil(0.90 × positive-image count)−1, with the implementation's fixed equality convention.

The operating selector compared no update, intercept, threshold and affine candidates on validation image loss |Se−0.90| + 0.1(1−Sp), retaining the specified tie order. Se and Sp denote image-level sensitivity and specificity. The probability selector compared no update, intercept and affine maps on validation Brier loss. When validation lacked either class, the selector retained no update. The published main comparison pairs the independently selected probability map with the selected operating cutoff. Its probabilities follow the Brier-selected map and its flags follow the operating selector.

TP and FN count recorded-positive patients with and without a positive flag, respectively; FP and TN count recorded-negative patients with and without a positive flag, respectively. Positive flags/1,000 = 1,000(TP+FP)/N. Missed positives/1,000 = 1,000 FN/N. Sensitivity = TP/(TP+FN), and specificity = TN/(TN+FP). Brier loss is the mean squared error over included images.

Net benefit, NB(p)=TP/N−FP/N × p/(1−p) at threshold probability p, holds each selected binary action fixed and weights false-positive flags against true-positive detections. The recorded grid was 0.05, 0.10, 0.20 and 0.30, displayed in full in the manuscript.

The added decision references use the same evaluation-patient denominators. Flag all gives sensitivity 1, specificity 0, 1,000 flags and zero missed positives per 1,000 patients. Flag none gives sensitivity 0, specificity 1, zero flags and 231.8653 missed positives per 1,000 in mBRSET or 498.3278 in DeepDRiD. Their net benefits are (q−p)/(1−p) and zero, respectively. Absolute strategy net benefit is reconstructed from TP/N=q−misses/1,000 and FP/N=flags/1,000−TP/N. The reconstruction agrees with the saved sensitivity/specificity and paired net-benefit summaries within 1e−9. The additional-flag ratio is (mean updated flags−mean unchanged flags)/(mean unchanged omissions−mean updated omissions), with a positive denominator required. It is a ratio of mean differences across the saved audits, rather than a mean of per-audit ratios. These additions were made after review of the existing results and are descriptive; the original intervals retain their original paired comparators. Full formulas and input hashes are supplied in evidence/decision_baseline_provenance.json.

### S4. COMPLETE PRIMARY RESULTS AND UNCERTAINTY

Supplementary table S1, supplied as source_tables/clinical_all_strategies_n200.csv, contains all seven named strategies for budget 200 across the four systems and two cohorts. It includes no update, fixed intercept, fixed affine, fixed threshold, operating selection, probability selection and the paired probability/operating controls. Some named actions have identical decisions.

Supplementary table S2, source_tables/clinical_primary_conditional_intervals.csv, contains the 24 source-specific primary rows, including Brier differences and net-benefit estimates and intervals at all four threshold probabilities. Each interval uses the existing 1,999 evaluation-patient cluster resamples and averages the saved actions within that source partition. It conditions on the source system, selected actions, observed patient pool and reference, so it excludes source-training and audit-selection uncertainty and is pointwise rather than simultaneous. Uncertainty is reported separately for each source partition; the three-partition mean is descriptive.

In mBRSET, the primary Brier decrease occurred in 147/150 APTOS audits, 139/150 ConvNeXt-T audits, 133/150 ViT-S16 audits and 143/150 RETFound audits. At p=0.20, concurrent Brier improvement and lower patient net benefit occurred in 141, 129, 114 and 104 of those dependent audits, respectively. These counts differ from the corresponding image-operating-loss discordance counts of 144, 139, 129 and 110 because Brier and the image operating criterion are distinct outcomes.

In DeepDRiD, Brier improved in 150, 150, 150 and 149 of 150 audits for APTOS, ConvNeXt-T, ViT-S16 and RETFound. Concurrent Brier improvement and lower net benefit at p=0.20 occurred in 0, 33, 82 and 104 audits. The favourable APTOS mean comparison and the heterogeneous foundation-system comparisons remain part of the study result.

All 24 source-specific Brier-difference intervals were below zero. At p=0.20, mBRSET had eight negative net-benefit intervals and four including zero; DeepDRiD APTOS had three positive intervals, while all other DeepDRiD intervals included zero. Restricting to fully graded mBRSET records retained negative p=0.20 intervals for both DINOv3 systems in all source partitions. The complete-grade data are supplied in source_tables/complete_grade_sensitivity.csv.

### S5. AUDIT SIZE, SELECTION BY PATIENT-LEVEL NET BENEFIT AND ALTERNATIVE OPERATING TARGETS

Budget-specific summaries and source-partition summaries are provided in source_tables/audit_budget_action_summary.csv and source_tables/audit_budget_split_summary.csv. The primary article foregrounds budget 200 to give a common supported comparison across systems. The smaller-budget summaries include no-update fallbacks in the original draw denominator; detailed candidate-level support and fallback states are retained in the original analysis records and are not included in these aggregate tables. A threshold may be unavailable because its specified positive-patient support is absent even when intercept or affine updates can be fitted.

The selector based on patient-level net benefit maximised validation patient-OR net benefit instead of the original image operating objective, using the same candidate family. Its protocol was written after net benefit at the four threshold probabilities had been inspected in these cohorts, so it is a development analysis. At budget 200 and p=0.20 in mBRSET, its mean improvements relative to the original selector were APTOS 0.0140, ConvNeXt-T 0.0520, ViT-S16 0.0388 and RETFound 0.0211. Its mean differences against no update were −0.0066, −0.0025, −0.0016 and −0.0038, and it retained no update in 69%, 95%, 96% and 65% of audits, respectively. In DeepDRiD at the same audit size and threshold probability, changes relative to the original selector were −0.0057, −0.0032, 0.0025 and −0.0009, and no update was retained in 8%, 35%, 61% and 27% of audits. source_tables/patient_aligned_selection_summary.csv retains all available audit sizes and threshold probabilities.

The alternative-target experiment used the two DINOv3 systems, two cohorts and three targets: sensitivity 0.85, sensitivity 0.95 and a source-specific specificity anchor. It retained its own complete-grade evaluation populations. At budget 200, fixed threshold adjustment had lower mean image operating loss than the selector in ten of twelve cells, tied in one and higher loss in one. All eight sensitivity-target cells favoured fixed threshold adjustment in this mean comparison; specificity anchoring supplied the exception. The twelve-cell table is supplied as source_tables/alternate_targets_n200.csv, with the saved independent arithmetic verification bound in the source manifest.

### S6. READOUT ADAPTATION AND RETENTION OF SOURCE SUPERVISION

Separate completed experiments compared fitting a new target-only logistic readout with retaining the source-supervised score as an offset and fitting an additional target correction. Their analysis population was defined by the original representation-specific ledgers for DINOv3 ViT-S16, ConvNeXt-T and RETFound, source partition 0, and the original patient audit/evaluation roles.

At budget 25, fixed-C=0.01 target-only replacement had negative difference intervals for patient-level area under the receiver operating characteristic curve (AUROC) against the frozen source in all three mBRSET representations and the two DeepDRiD DINOv3 systems; DeepDRiD RETFound's interval included zero. A fit-only patient cross-validation choice of C retained this five-negative/one-including-zero pattern. Retaining the source score improved patient AUROC over the cross-validation-selected replacement in all six cohort–representation comparisons, while comparisons against the frozen source were mixed. The 24 relevant completed interval rows are supplied in source_tables/readout_adaptation_n25_ranking.csv; its analysis and comparator columns distinguish these controls.

At budget 200, retaining the source score produced positive AUROC-difference intervals against the frozen source in five of six comparisons. Against fixed-C target-only replacement, two intervals were above zero and four included zero. At an audit flagging fraction request of 0.50, omission-difference intervals against source thresholds set using all 200 audit patients were negative in five comparisons and included zero in one. Flagging-difference intervals were positive in four and included zero in two. The full paired table retains source thresholds estimated from the 60-patient validation subset and from the full 200-patient audit, target-only replacement and the frozen source as distinct comparators.

The flagging-fraction request was applied within the audit; paired differences in realised evaluation flagging rates are reported against the named comparators. The controls retain their specified supervision, regularisation and stopping rules. Their patient/draw bootstrap intervals condition on the saved fits and evaluation pools. The full budget-200 ranking and operating tables are supplied as source_tables/source_retention_ranking_intervals.csv and source_tables/source_retention_operating_intervals.csv. methods/supplementary_evidence_index.pdf identifies the exact row filters, source files and hashes.

### S7. REFERENCE AND INPUT-COVERAGE CONTEXT

The completed BRSET released-reference comparison held images and predictions fixed while substituting grades from the earlier v1.0.0 and v1.0.1 releases; all primary analyses used v1.0.2 grades. For grade ≥3, 124 recorded positives became negatives and 14 negatives became positives. Sensitivity differences for ViT-S16 and ConvNeXt-T were +0.0360 and +0.0381, with positive predictive value differences −0.0703 and −0.0492, respectively (source_tables/brset_reference_revision_fixed_predictions.csv). These are a recorded reference-version contrast rather than two independent cohorts. The operating selector's chosen action changed in 12–48 of 100 dependent audits across four representation–endpoint settings at budget 200. Under each evaluation reference, mean loss-difference signs against no update agreed between old-reference and new-reference selection (source_tables/brset_cross_reference_policy_n200.csv).

The separate mBRSET image/reference coverage analysis used 1,094 patients with complete four-image support, divided into 444 audit and 650 evaluation patients. Recorded two-image reference subsets omitted 11–15 of the full-reference positive evaluation patients (source_tables/mbrset_reference_coverage_omitted_counts.csv). Crossing full/subset inputs with full/subset recorded references produced 48 aggregate cells, supplied in source_tables/mbrset_input_reference_coverage_cells.csv. These show that changing input coverage and changing reference coverage could move measured discrimination in different directions. This population is distinct from the primary 772-patient evaluation pool. A maintenance report should therefore state which images and which reference define each patient's result. Source files and population provenance are listed in the supplementary evidence index.

### S8. OTHER COMPLETED ANALYSES AND THE SCOPE OF THIS ARTICLE

The wider research programme included MMRDR, IDRiD, DDR, MSHF, HYAMD, OLIVES, MARIO and EyePACS analyses addressing acquisition, reference structure, image quality, other disease endpoints or OCT tasks. This article brings together the patient-linked diabetic-retinopathy comparisons and controls relevant to detection and flagging after local updating.

### S9. REPRODUCIBILITY MATERIALS

The source manifest links publication tables and figures to the original aggregate files, analysis code and saved arithmetic checks. The accompanying code reproduces publication tables and figures from the supplied aggregates. Source code is supplied for the primary four-system replay, its patient-cluster bootstrap, the patient-level selector and the APTOS maintenance analysis; the scripts that produced the S5 alternative-target, S6 readout and S7 reference-coverage aggregates, and the feature-extraction and inference workers, are not included; their result files are identified in the methods attachments. methods/supplementary_evidence_index.pdf gives the packaged S6/S7 tables, row filters, definitions and source hashes. Original model inference, head fitting and patient-level bootstrap records require the authorised original inputs and saved states listed in section 7 of methods/model_provenance_and_locked_methods.pdf. The archive contains aggregate results and selected analysis sources; original images, weights and individual-level records remain subject to provider access terms.

### SUPPLEMENTARY DISPLAYS

**Supplementary table S2. Source-specific Brier and net-benefit differences with existing 95% conditional patient-cluster intervals.**

| **Cohort / system** | **Source partition** | **ΔBrier [95% CI]** | **ΔNB at p=0.20 [95% CI]** |
| --- | --- | --- | --- |
| mBRSET / APTOS | 0 | -0.0509 [-0.0645, -0.0369] | -0.0287 [-0.0560, -0.0007] |
| mBRSET / APTOS | 1 | -0.0528 [-0.0664, -0.0398] | -0.0183 [-0.0459, 0.0110] |
| mBRSET / APTOS | 2 | -0.0492 [-0.0636, -0.0345] | -0.0149 [-0.0402, 0.0099] |
| mBRSET / DINO-Conv | 0 | -0.0349 [-0.0459, -0.0251] | -0.0605 [-0.0809, -0.0394] |
| mBRSET / DINO-Conv | 1 | -0.0251 [-0.0333, -0.0169] | -0.0435 [-0.0582, -0.0284] |
| mBRSET / DINO-Conv | 2 | -0.0241 [-0.0321, -0.0165] | -0.0595 [-0.0764, -0.0423] |
| mBRSET / DINO-ViT | 0 | -0.0143 [-0.0205, -0.0087] | -0.0226 [-0.0341, -0.0106] |
| mBRSET / DINO-ViT | 1 | -0.0169 [-0.0241, -0.0100] | -0.0415 [-0.0572, -0.0241] |
| mBRSET / DINO-ViT | 2 | -0.0136 [-0.0193, -0.0084] | -0.0570 [-0.0739, -0.0387] |
| mBRSET / RETFound | 0 | -0.0388 [-0.0518, -0.0269] | 0.0021 [-0.0323, 0.0345] |
| mBRSET / RETFound | 1 | -0.0324 [-0.0432, -0.0222] | -0.0488 [-0.0716, -0.0240] |
| mBRSET / RETFound | 2 | -0.0271 [-0.0365, -0.0185] | -0.0278 [-0.0525, 0.0001] |
| DeepDRiD / APTOS | 0 | -0.2249 [-0.2656, -0.1831] | 0.0426 [0.0036, 0.0843] |
| DeepDRiD / APTOS | 1 | -0.2050 [-0.2439, -0.1660] | 0.0686 [0.0278, 0.1116] |
| DeepDRiD / APTOS | 2 | -0.2101 [-0.2471, -0.1715] | 0.0418 [0.0038, 0.0818] |
| DeepDRiD / DINO-Conv | 0 | -0.1733 [-0.2074, -0.1398] | 0.0005 [-0.0298, 0.0340] |
| DeepDRiD / DINO-Conv | 1 | -0.1548 [-0.1893, -0.1218] | 0.0032 [-0.0244, 0.0312] |
| DeepDRiD / DINO-Conv | 2 | -0.1610 [-0.2000, -0.1262] | 0.0079 [-0.0260, 0.0467] |
| DeepDRiD / DINO-ViT | 0 | -0.1022 [-0.1293, -0.0764] | -0.0076 [-0.0234, 0.0109] |
| DeepDRiD / DINO-ViT | 1 | -0.0936 [-0.1198, -0.0674] | -0.0022 [-0.0142, 0.0111] |
| DeepDRiD / DINO-ViT | 2 | -0.0939 [-0.1203, -0.0685] | -0.0026 [-0.0235, 0.0220] |
| DeepDRiD / RETFound | 0 | -0.1110 [-0.1383, -0.0844] | 0.0073 [-0.0203, 0.0383] |
| DeepDRiD / RETFound | 1 | -0.1180 [-0.1506, -0.0868] | -0.0082 [-0.0324, 0.0178] |
| DeepDRiD / RETFound | 2 | -0.1061 [-0.1336, -0.0771] | -0.0122 [-0.0364, 0.0138] |

Each row uses 50 fixed selected audits and 1,999 evaluation-patient cluster resamples. The 24 rows are dependent across partitions and systems. The downloadable CSV also includes p=0.05, 0.10 and 0.30.

**Figure S1. Audit budget and additional positive flags.** Means across three source partitions and 50 audits per budget are displayed for the primary selected operating policy.

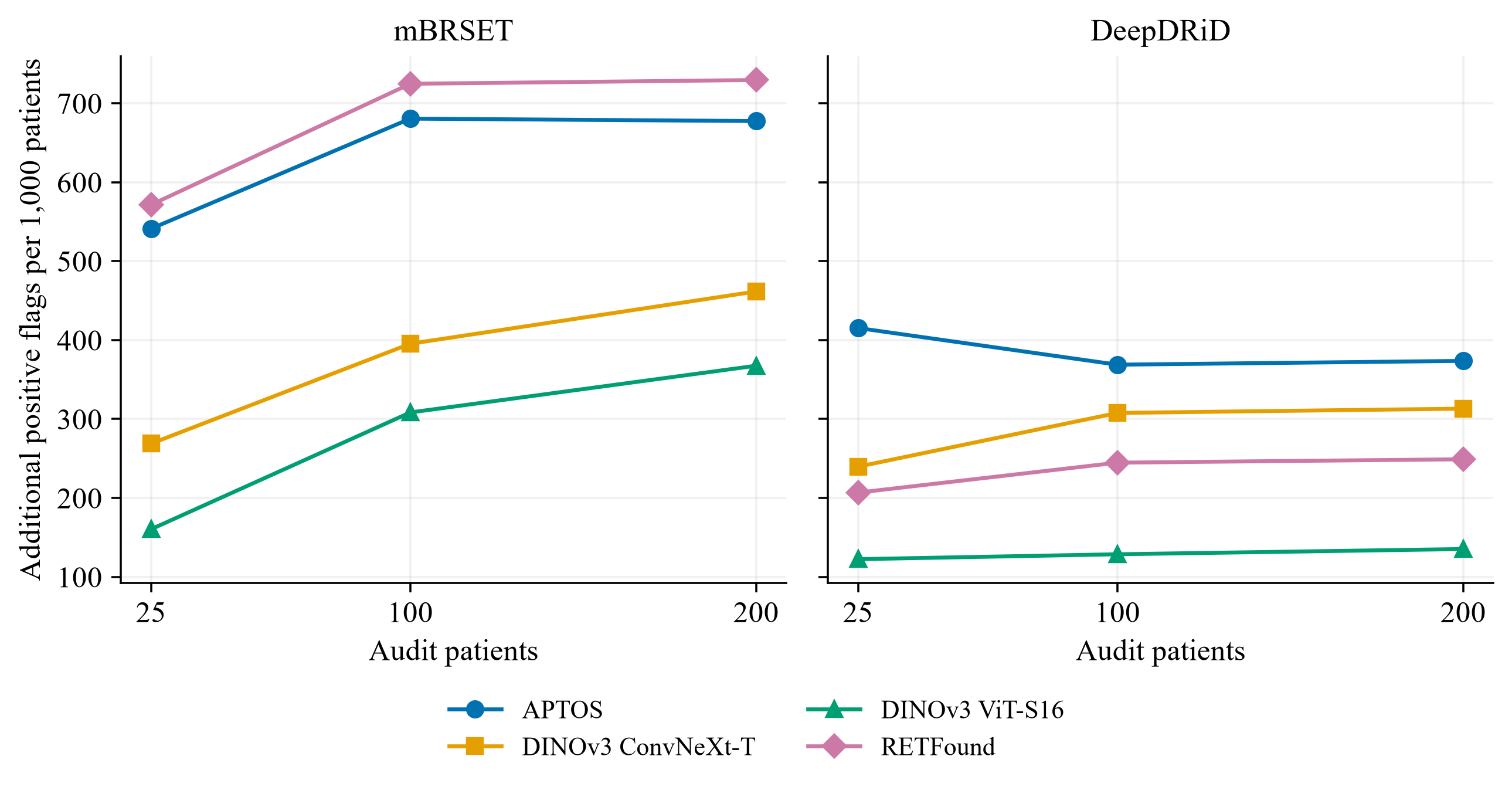

**Figure S2. Selection by patient-level net benefit.** Mean incremental net benefit at budget 200 and p=0.20 compares selection by patient-level net benefit with the original image-based selector and with no update.

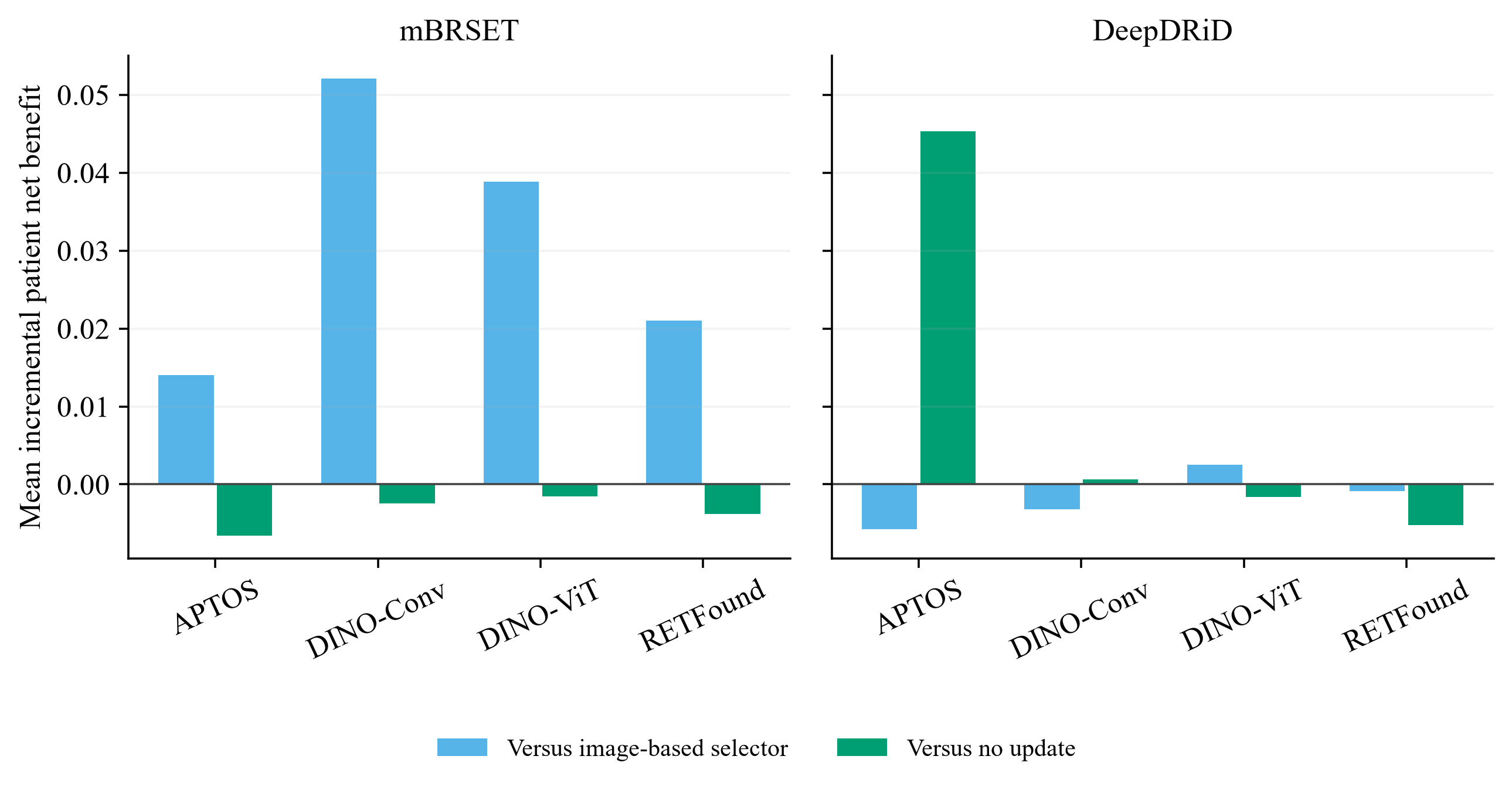

**Supplementary table S3. Descriptive decision references and additional-flag ratios at budget 200.**

| **Cohort / system** | **Extra flags / extra detection** | **Selected NB, p=0.05** | **ΔNB vs flag all, p=0.05** | **Selected NB, p=0.20** | **ΔNB vs flag all, p=0.20** |
| --- | --- | --- | --- | --- | --- |
| mBRSET / APTOS | 5.69 | 0.1860 | -0.0054 | 0.0651 | +0.0252 |
| mBRSET / DINO-Conv | 9.48 | 0.1905 | -0.0009 | 0.0602 | +0.0204 |
| mBRSET / DINO-ViT | 8.92 | 0.1908 | -0.0007 | 0.0748 | +0.0350 |
| mBRSET / RETFound | 5.79 | 0.1917 | +0.0002 | 0.0493 | +0.0095 |
| DeepDRiD / APTOS | 3.23 | 0.4644 | -0.0075 | 0.4014 | +0.0284 |
| DeepDRiD / DINO-Conv | 4.77 | 0.4770 | +0.0050 | 0.4055 | +0.0326 |
| DeepDRiD / DINO-ViT | 5.70 | 0.4737 | +0.0017 | 0.4175 | +0.0446 |
| DeepDRiD / RETFound | 5.38 | 0.4700 | -0.0020 | 0.3947 | +0.0218 |

Flag-none net benefit is zero. Flag-all net benefit at p=0.05/0.20 is 0.1914/0.0398 in mBRSET and 0.4719/0.3729 in DeepDRiD. The ratio divides the mean additional flags by the mean reduction in missed positive patients, including the newly detected patients among additional flags. All ratios and new flag-all/flag-none comparisons are descriptive. Full precision, all four threshold probabilities and all seven strategy labels are supplied in source_tables/decision_reference_baselines_n200.csv and source_tables/additional_flags_per_detection_n200.csv; selection by patient-level net benefit is supplied in source_tables/patient_aligned_decision_baselines.csv.
